# SARS-CoV-2 variant of concern B.1.1.7: diagnostic accuracy of three antigen-detecting rapid tests

**DOI:** 10.1101/2021.06.15.21258502

**Authors:** Andreas K. Lindner, Lisa J. Krüger, Olga Nikolai, Julian A.F. Klein, Heike Rössig, Paul Schnitzler, Victor M. Corman, Terry C. Jones, Frank Tobian, Mary Gaeddert, Susen Burock, Jilian A. Sacks, Joachim Seybold, Frank P. Mockenhaupt, Claudia M. Denkinger

## Abstract

Virus mutations have the potential to impact the accuracy of diagnostic tests. The SARS-CoV-2 B.1.1.7 lineage is defined by a large number of mutations in the spike gene and four in the nucleocapsid (N) gene. Most commercially available SARS-CoV-2 antigen-detecting rapid tests (Ag-RDTs) target the viral N-protein, encoded by the N-gene.

We conducted a manufacturer-independent, prospective diagnostic accuracy study of three SARS-CoV-2 Ag-RDTs that are currently under review by the WHO Emergency Use Listing Procedure (Espline -Fujirebio Inc.; Sure Status -Premier Medical Corporation Private Limited; Mologic -Mologic Ltd.) and report here on an additional sub-analysis regarding the B.1.1.7 lineage. During the study, in Berlin and Heidelberg, Germany, from 20 January to 15 April 2021, B.1.1.7 rapidly became the dominant SARS-CoV-2 lineage at the testing sites and was detected in 220 (62%) of SARS-CoV-2 RT-PCR positive patients. All three Ag-RDTs yielded comparable sensitivities irrespective of an infection with the B.1.1.7 lineage or not.

There is only limited data on how N-gene mutations in variants of concern may impact Ag-RDTs. Currently, no major changes to test performance are anticipated. However, test developers and health authorities should assess and monitor the impact of emerging variants on the accuracy of Ag-RDTs.

## Manuscript

The SARS-CoV-2 B.1.1.7 lineage (recently labelled as Alpha by WHO), also known as variant of concern (VOC) 202012/01, rapidly emerged after its first identification in the United Kingdom in late 2020 (1). As of 1 June 2021, B.1.1.7 has been verified in 160 countries and become the dominant variant in several European countries, including Germany, and in North America (2-4). B.1.1.7 is defined by a large number of mutations in the spike (S) gene and four in the nucleocapsid (N) gene (N-protein: D3L, R203K, G204R, and S235F) (5). Virus mutations have the potential to impact the accuracy of diagnostic tests. Most commercially available SARS-CoV-2 antigen-detecting rapid tests (Ag-RDTs) target the C-terminus of the viral N-protein, encoded by the N-gene (6, 7). The N-gene mutations of B.1.1.7 are located at the N-terminus (7). In an evaluation by Public Health England, the B.1.1.7 variant did not affect the performance of six commercially available Ag-RDTs, despite a limited number of amino acid changes from the original viral sequence in the target antigen (5, 8).

We conducted a manufacturer-independent, prospective diagnostic accuracy study of three SARS-CoV-2 Ag-RDTs at an ambulatory testing facility in Berlin and Heidelberg, Germany, from 20 January to 15 April 2021. All three Ag-RDTs evaluated are currently under review by the WHO Emergency Use Listing Procedure: 1) Espline SARS-CoV-2 (Fujirebio Inc.); 2) Sure Status COVID-19 Antigen Card Test (Premier Medical Corporation Private Limited), both using nasopharyngeal sampling; and 3) Mologic COVID-19 Rapid Test (Mologic Ltd.), using anterior nasal sampling (9). Reference standard was RT-PCR using naso-/oropharyngeal sampling. Study procedures were previously described (10). The study was conducted in collaboration with the Foundation of Innovative New Diagnostics (FIND), a WHO Collaborating Centre. Here we report on an additional sub-analysis regarding the B.1.1.7 lineage.

Of 1692 adults enrolled in the study, 354 (21%) tested positive by RT-PCR. Positive samples were typed for the N501Y and del69–70 polymorphisms by melting curve analysis. The presence of both polymorphisms was considered indicative of a B.1.1.7 lineage infection, an inference whose accuracy was confirmed by full-genome sequencing of all Heidelberg samples. During the study, B.1.1.7 became the dominant SARS-CoV-2 lineage within only five weeks (11). Among the positive patients, 220 (62%) were infected with B.1.1.7, 3 (1%) with VOC B.1.351 and 109 (31%) with wildtype virus or mutations that are currently considered not of concern. All three Ag-RDTs yielded comparable sensitivities irrespective of an infection with the B.1.1.7 lineage or not (Table 1). In 22 (6%) patients, typing was not done or not possible due to the limit of detection of genotyping for samples with low viral load (excluded from analysis and leading to a slight overestimation of sensitivity).

**TABLE 1.**
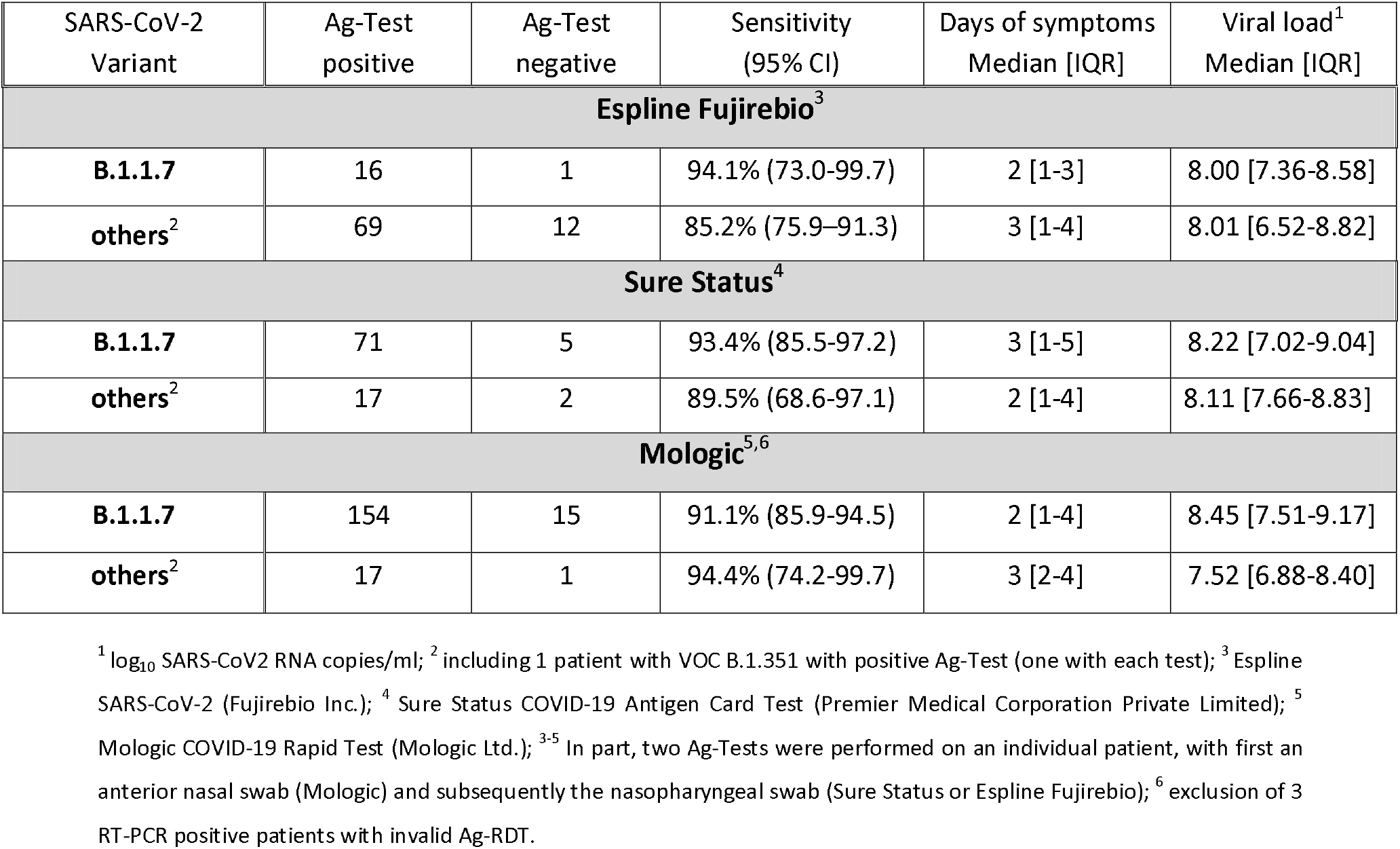
Results of three antigen rapid tests with sensitivity, median duration of symptoms, and median viral load for 329 RT-PCR positive patients with viral typing. Results from patients infected with B.1.1.7 are shown separately from others (i.e., infected with wildtype virus or other mutations). Sensitivities are slightly overestimated due to the limit of detection of genotyping for samples with low viral load.

| SARS-CoV-2 Variant | Ag-Test positive | Ag-Test negative | Sensitivity (95% CI) | Days of symptoms Median [IQR] | Viral load<sup>1</sup> Median [IQR] |
| --- | --- | --- | --- | --- | --- |
| **Espline Fujirebio<sup>3</sup>** |  |  |  |  |  |
| **B.1.1.7** | 16 | 1 | 94.1% (73.0-99.7) | 2 [1-3] | 8.00 [7.36-8.58] |
| **others<sup>2</sup>** | 69 | 12 | 85.2% (75.9-91.3) | 3 [1-4] | 8.01 [6.52-8.82] |
| **Sure Status<sup>4</sup>** |  |  |  |  |  |
| **B.1.1.7** | 71 | 5 | 93.4% (85.5-97.2) | 3 [1-5] | 8.22 [7.02-9.04] |
| **others<sup>2</sup>** | 17 | 2 | 89.5% (68.6-97.1) | 2 [1-4] | 8.11 [7.66-8.83] |
| **Mologic<sup>5,6</sup>** |  |  |  |  |  |
| **B.1.1.7** | 154 | 15 | 91.1% (85.9-94.5) | 2 [1-4] | 8.45 [7.51-9.17] |
| **others<sup>2</sup>** | 17 | 1 | 94.4% (74.2-99.7) | 3 [2-4] | 7.52 [6.88-8.40] |

<sup>1</sup> log<sub>10</sub> SARS-CoV2 RNA copies/ml; <sup>2</sup> including 1 patient with VOC B.1.351 with positive Ag-Test (one with each test); <sup>3</sup> Espline SARS-CoV-2 (Fujirebio Inc.); <sup>4</sup> Sure Status COVID-19 Antigen Card Test (Premier Medical Corporation Private Limited); <sup>5</sup> Mologic COVID-19 Rapid Test (Mologic Ltd.); <sup>3-5</sup> In part, two Ag-Tests were performed on an individual patient, with first an anterior nasal swab (Mologic) and subsequently the nasopharyngeal swab (Sure Status or Espline Fujirebio); <sup>6</sup> exclusion of 3 RT-PCR positive patients with invalid Ag-RDT.

There is only limited data on how N-gene mutations in VOCs may impact Ag-RDTs and this clinical evaluation complements analytical evaluations (5, 8, 12, 13). Currently, no major changes to test performance are anticipated (6). However, test developers and health authorities should assess and monitor the impact of emerging variants on Ag-RDTs during development and post-authorization (14).

## Data Availability

All raw data and analysis code are available upon a request to the corresponding author.

## Author contributions

AKL: conceptualization, methodology, writing -original draft. LJK: investigation, formal analysis. ON: methodology, investigation, formal analysis. JAFK: investigation, formal analysis. HR: project administration. PS: investigation, formal analysis. VMC: investigation, formal analysis. TCJ: investigation, formal analysis. FT: formal analysis. MG: formal analysis. SB: project administration. JAS: supervision. JS: supervision, project administration. FPM: conceptualization, supervision. CMD: conceptualization, methodology, supervision. All authors: writing -review & editing.

## Acknowledgements

Chiara Rohardt, Maximilian Gertler, Elisabeth Linzbach, Domenika Pettenkofer, Christian Schönfeld, Julian Bernhard.

## Data availability

All raw data and analysis code are available upon a request to the corresponding author.

## Ethics

This study was approved by the ethics committee of Charité -Universitätsmedizin (EA1/371/20) and of the Heidelberg University Hospital (Registration number S-180/2020).

